# Prospective follow-up of New York City residents with e-cigarette, or vaping product use-associated lung injury—2020–2021

**DOI:** 10.1101/2024.05.23.24307782

**Authors:** Kathryn M. Tannert Niang, Aviva B. Grasso, Indira Debchoudhury, Dena Bushman, John P. Jasek, Monique A. Fairclough, Katherine R. Van Oss, Shadi Chamany, Kendall D. LaSane, Sharraine M. Franklin, Achala K. Talati

**Author notes:** Corresponding author (KTN). The findings and conclusions in this report are those of the authors and do not necessarily represent the official position of the Centers for Disease Control and Prevention.

## Abstract

**Background:** A multistate outbreak of e-cigarette, or vaping, product use-associated lung injury (EVALI) occurred in 2019. Because of EVALI’s novelty and severity, the New York City (NYC) Department of Health and Mental Hygiene (DOHMH) prospectively assessed sequelae among NYC residents who received an EVALI diagnosis in 2019.

**Methods:** Using existing NYC EVALI surveillance data, DOHMH attempted contact with all living residents who received an EVALI diagnosis in 2019 and conducted 3 waves of telephone interviews during April 2020–March 2021. Interview questions were adapted from the Centers for Disease Control and Prevention’s EVALI case report form and validated surveys. Baseline differences between respondents and nonrespondents were assessed with Chi-square and Fisher’s exact tests; clinical and behavioral characteristics and open-ended responses were summarized.

**Results:** In 2019, 53 NYC residents received an EVALI diagnosis; 33 (67%), 14 (29%), and 18 (37%) of 49 living residents participated in the first, second, and third interviews, respectively. Interviews occurred after outpatient diagnosis (6%) or hospital discharge (94%), at a median of 8, 11, and 17 months for each wave. Respondents (N = 33) and nonrespondents (N = 16) did not differ by sex, age, hospitalization status or length. Respondents were mostly male (70%), had a median age of 23 years (range: 16–63 years), and all reported using vaping or e-cigarette products (vaping) with tetrahydrocannabinol (88%), nicotine (49%), or cannabidiol (9%) before diagnosis. Respiratory (first and second interviews) and gastrointestinal (third interviews) symptoms were most commonly reported. Sixteen respondents (49%) reported any new diagnosis during follow-up. Fifteen to 29% of respondents reported vaping at each interview; 58%–93% reported recent non-vaped cannabinoid use.

**Conclusion:** NYC residents with EVALI reported symptoms throughout the follow-up period, and approximately half reported newly diagnosed health conditions.

## Introduction

In summer 2019, the Centers for Disease Control and Prevention (CDC), the Food and Drug Administration, and state and local health departments, including New York City (NYC) Department of Health and Mental Hygiene (DOHMH), began investigating reports of severe lung injuries among persons with a history of e-cigarette or vaping product use (vaping). During August 2019–February 2020, CDC reported 2,807 cases of e-cigarette, or vaping, product use-associated lung injury (EVALI) nationwide. This included 127 (5%) that were reported in New York State (NYS) during August 2019-February 2020.[1] During August–December 2019, NYS reported 113 cases, of which 53 (46%) were NYC residents and are the subject of this report.

CDC described patients from across the United States who received an EVALI diagnosis mostly were young adult males. These persons had vaped tetrahydrocannabinol (THC), nicotine, or other liquids (e-liquids), and experienced respiratory, gastrointestinal, and constitutional symptoms over the course of a few days to several weeks [1]. Because of reports of rehospitalization and death post-discharge from initial EVALI hospitalization, CDC’s guidance on standards of care evolved between the initial recognition of EVALI in August 2019, and January 2020, when management algorithms were updated. This guidance addressed patients’ clinical readiness for hospital discharge, social support and access to mental health and substance use disorder services, best practices for medication adherence, and post-discharge medical follow-up, including with primary care providers and pulmonologists [2].

Despite the clinical severity of certain EVALI cases and concerns about the toxicity of vaping products more broadly,[3, 4] the long-term effects of EVALI are not well understood [2, 5, 6]. Because of the novelty of EVALI and reports of rehospitalization and death, DOHMH prospectively assessed NYC residents who had received an EVALI diagnosis in 2019. Even with limited knowledge on sequelae of EVALI, we were able to assess changes in symptoms, concomitant diagnoses, healthcare usage, functional and financial consequences, and substance use over time, among this cohort of NYC residents.

## Methods

### Data Sources

In 2019, potential EVALI cases were identified through passive (reports to poison control centers, NYS Department of Health [DOH], and DOHMH) and active (syndromic) surveillance. At that time, each potential EVALI case was identified and classified using CDC’s 2019 Lung Injury surveillance case definitions [7]. Baseline EVALI surveillance data were collected from provider reports to the NYC Poison Control Center and patient medical records that were obtained directly from NYC hospitals. During April 2020–March 2021, we conducted 3 waves of telephone interviews with surviving NYC residents who had received an EVALI diagnosis in 2019 to follow them over time; the list was confirmed with NYS DOH to ensure that all reported cases were included. Before each wave of interviews, a match with the NYC vital statistics death registry was conducted to assess for any recent deaths.

We conducted interviews in English or in respondent’s primary language (n = 1, in Mandarin) using a telephone interpreter service. NYC residents who received an EVALI diagnosis were called up to three times during each wave of interviews. For those not reached in the first call attempt, we left voicemails and sent text messages or emails to help schedule interviews. We mailed letters to all persons not reached during the first interview wave, informing them of the surveillance project and requesting their participation. Approximately three months after completing the first wave of interviews, we initiated second interviews with the initial respondents. We also attempted to interview all nonrespondents from the first wave of calls who had not opted out of participation, completing abbreviated first interviews with those reached. After another three months, the final wave of interviews began, including only respondents with whom at least one previous interview had been completed (Fig 1).

**Fig 1:**
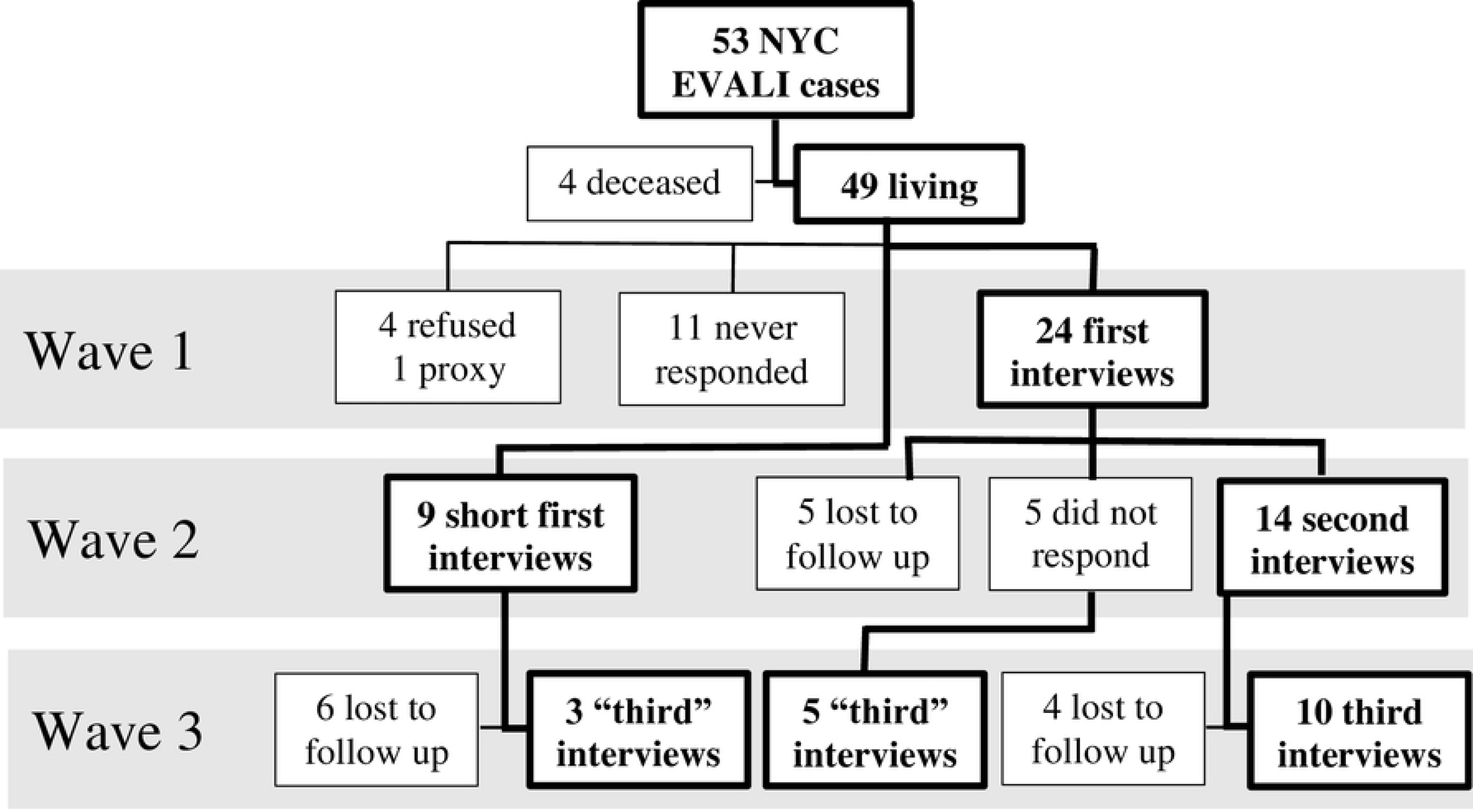
Enrollment and participation in NYC e-cigarette, or vaping, associated lung injury (EVALI) interviews, waves 1–3.

Interview questions included age, sex assigned at birth, symptoms, health conditions, healthcare use, insurance coverage, substance use, functional, student, and employment status. Effects of EVALI on finances were mostly categorical and adapted from CDC’s EVALI case report form [8] and validated surveys. This included the NYC Community Health Survey, [9] Youth Risk Behavior Survey, [10] and National Survey on Drug Use and Health [11]. Abbreviated interviews included a shorter set of primarily open-ended questions that covered the same content areas. At the end of each interview, we asked if respondents had anything else to share about their experience or consequences of their lung injury on their lives.

Information about hospitalization, including duration of stay and intubation status, were abstracted from medical records, along with sex, age, and race and ethnicity. We did not include interview questions about race and ethnicity, because we had intended to use NYS DOH case interview data from the initial outbreak. However, these data were missing for most NYC residents with an EVALI diagnosis, something we did not learn until the follow-up interviews were already completed. Instead, we used electronic medical record data to supplement this information because provider reports, which were used to identify cases did not include race or ethnicity.

Data were stored in restricted access file folders on secure agency share drives. This activity was reviewed by CDC and DOHMH Institutional Review Boards and was determined to be non-research public health surveillance. This study was reviewed by CDC and was conducted consistent with federal law and CDC policy.^1^

### Data analyses

Data from the first wave of interviews using the full interview form and data from those interviewed for the first time during the second wave using the abbreviated interview form, were merged and recoded as first interview. Open-ended responses from the abbreviated interviews were coded to match the categories from the full interview form, when possible. All three waves of interviews were coded as third interview, regardless of the number of prior interviews.

Baseline differences between respondents and nonrespondents were assessed with Chi-square and Fisher’s exact tests using SAS^®^ version 9.4 (SAS Institute, Inc., Cary, North Carolina).

Symptoms, health conditions, healthcare use, substance use patterns, functional status, and effects of EVALI on finances were also summarized for each wave of interviews. Symptom trends among the respondents who completed all three interviews were summarized to assess for resolution over time. Two investigators reviewed free text comments for additional themes and to provide context for the quantitative data. Because of missing data, race and ethnicity were not used in any analyses.

## Results

### Baseline characteristics

In 2019, a total of 53 NYC residents received an EVALI diagnosis. Most were male (72%); median age was 23 years (range: 15–63 years). Four (8%) died of EVALI (Table 1). Decedents were mostly male (n = 3; 75%) and had a median age of 28 years (range: 17–34 years). Two decedents had been hospitalized (mean = 13 days) and intubated; one decedent died at home, and one decedent presented to the emergency department in cardiac arrest and did not survive.

**Table 1:**
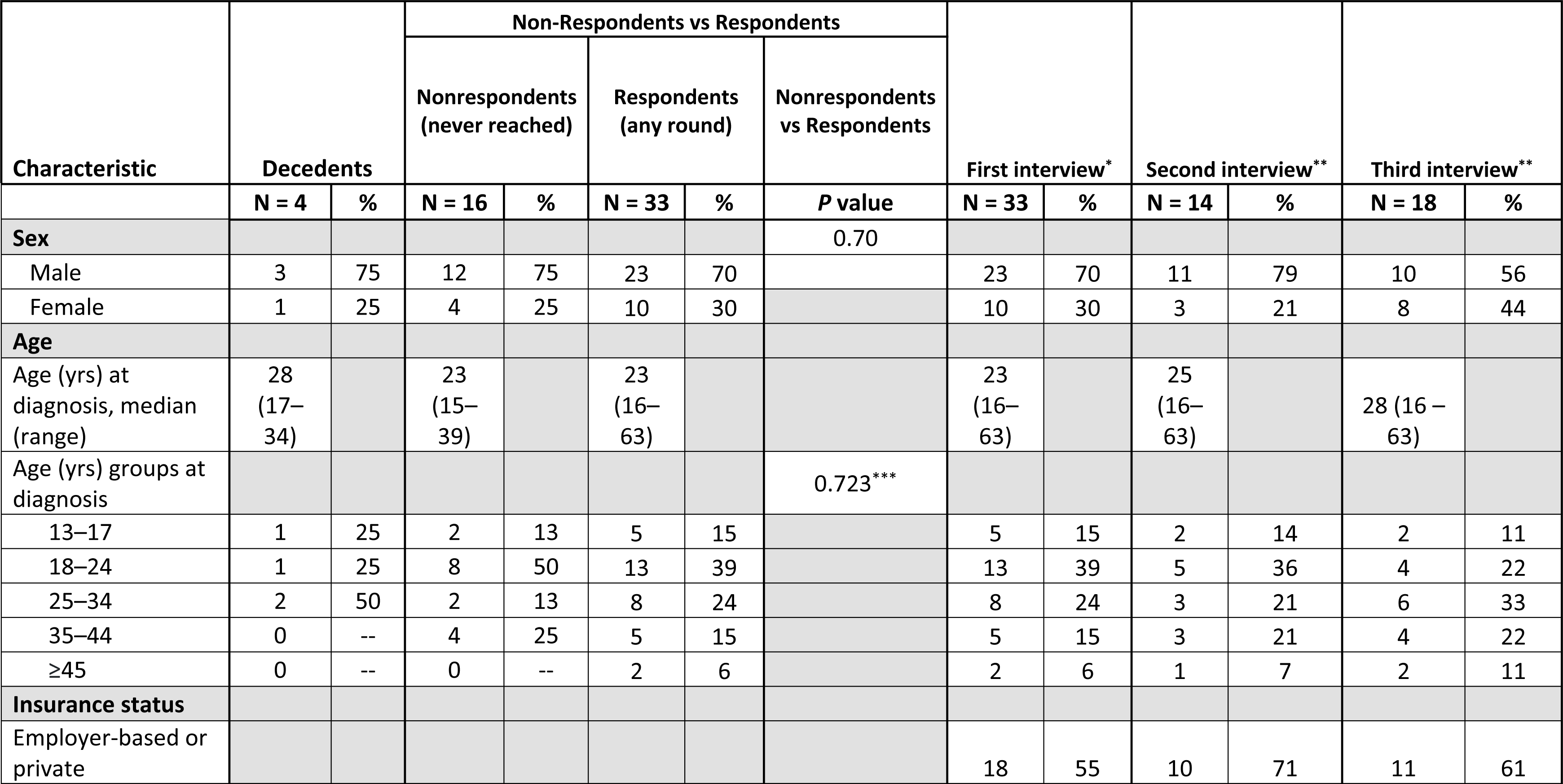
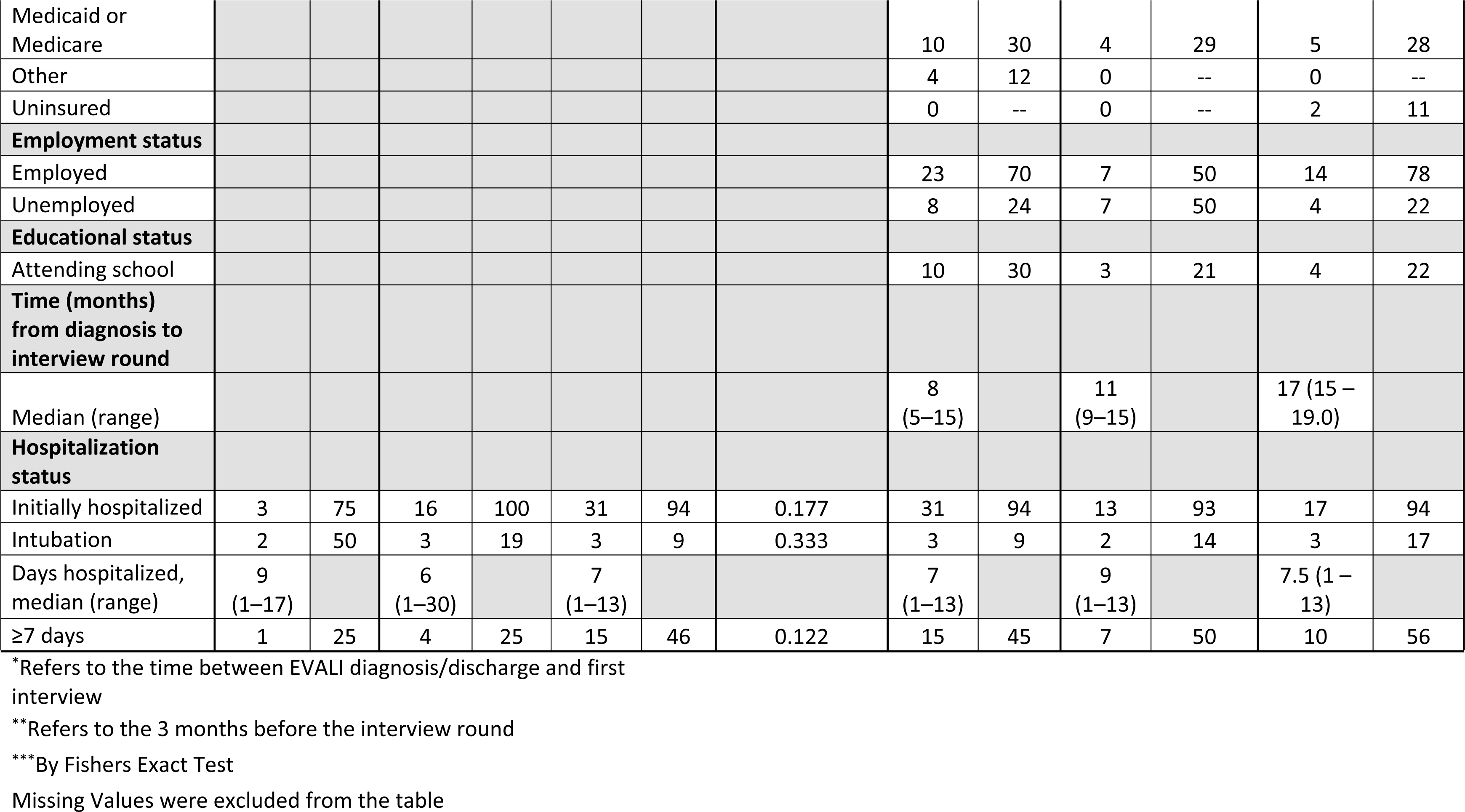
Demographic characteristics and hospitalization status among New York City residents who received a diagnosis of e-cigarette, or vaping, associated lung injury (EVALI) in 2019.

### Interview participation and respondent demographics

Interviews occurred from 5 to 19 months after initial hospitalization or injury, at a median of 8, 11, and 17 months for each interview wave. Among 49 potential respondents, 33 (67%) completed the first interview (24 in the first wave and 9 during the second wave), 14 (29%) completed the second interview, and 18 (37%) completed the third interview; overall, 33 (67%) respondents completed ≥1 interview and 22 (45%) completed ≥2 interviews. Ten respondents (20%) completed interviews during all three waves (Fig 1).

Respondents (N = 33; 67%) and nonrespondents (N = 16; 33%) did not significantly differ by sex, age group at diagnosis, intubation status, hospitalization status, or length of hospitalization (Table 1). Respondents were mostly male (n = 23; 70%), had a median age of 23 years (range: 16–63 years), were hospitalized (n = 31; 94%) for a median of 6 days (range: 0–13 days), and were not intubated (n = 30; 91%). Approximately half of respondents reported that at the time of hospitalization or diagnosis they had employer-based or private health insurance (n = 18; 55%). Race and ethnicity were incomplete for 16 of 49 surviving NYC residents who received an EVALI diagnosis in 2019 (33% of potential respondents) and data were not used.

In terms of substance use patterns, 100% of respondents reported vaping, with either tetrahydrocannabinol (THC) (88%), nicotine (49%), or cannabidiol (CBD) (9%) before diagnosis (Table 2). Some respondents (15%–29%) reported recent vaping at each interview, and most respondents (58%–93%) reported recent non-vaped (e.g., smoked or edible) cannabinoid use. A limited number of respondents reported smoking cigarettes (0%–6%) or other tobacco products (0%–11%).

**Table 2:** Vaping, e-cigarette, and substance use among New York City e-cigarette, or vaping, associated lung injury (EVALI) prospective follow-up respondents.

| Characteristic | Prior to EVALI diagnosis |  | First interview* |  | Second interview** |  | Third interview** |  |
| --- | --- | --- | --- | --- | --- | --- | --- | --- |
|  | N = 33 | % | N = 33 | % | N = 14 | % | N = 18 | % |
| **Vaped or used e-cigarettes** | 33 | 100 | 5 | 15 | 4 | 29 | 3 | 17 |
| Nicotine | 16 | 49 | 6 | 18 | 2 | 14 | 2 | 11 |
| Marijuana or THC | 29 | 88 | 2 | 6 | 2 | 14 | 2 | 11 |
| CBD | 3 | 9 | 1 | 3 | 0 | -- | 0 | -- |
| **General substance use** |  |  |  |  |  |  |  |  |
| Cigarettes |  |  | 2 | 6 | 0 | -- | 1 | 6 |
| Other tobacco products |  |  | 2 | 6 | 0 | -- | 2 | 11 |
| Hookah or Waterpipe |  |  | 1 | 3 | 1 | 7 | 1 | 6 |
| Non-Vaped Cannabinoids |  |  | 19 | 58 | 13 | 93 | 13 | 72 |
| Smoked or Dabbed |  |  |  |  | 9 | 64 | 7 | 39 |
| Other substance (heroin; cocaine; methamphetamines; huffing [paint or glue]; other) |  |  | 2 | 6 | 0 | -- | 4 | 22 |
| **Alcohol use** |  |  |  |  |  |  |  |  |
| ≥ 1 drink in last 30 days |  |  |  |  | 9 | 64 | 12 | 67 |
| Binge drank in last 30 days<sup>1</sup> |  |  |  |  | 1 | 7 | 3 | 17 |

\*Refers to the time between EVALI diagnosis and discharge and first interview.
\*\*Refers to the 3-month period before the interview.
<sup>1</sup>Binge refers to ≥4 drinks in a row for females, ≥5 drinks in a row for males.
Missing values were excluded from the table.

### Preexisting and Newly Diagnosed Health Conditions

Respondents reported preexisting mental health conditions (49%), including anxiety (36%), depression (24%) and other conditions (15%); asthma (27%), cardiovascular (12%) conditions, including hypertension, hyperlipidemia, and heart disease; and diabetes (3%). At first interview, respondents reported receiving new diagnoses of anxiety (27%), depression (15%), other mental health conditions (3%), asthma (9%), other respiratory (3%) or cardiovascular (9%) conditions, and diabetes (3%) (Table 3). Overall, 16 respondents (49%) reported receiving a new diagnosis during the follow-up period.

**Table 3:** Clinical characteristics and effects of e-cigarette, or vaping, associated lung injury (EVALI) diagnosis among respondents.

| Characteristic | Before EVALI diagnosis |  | First interview* |  | Second interview** |  | Third interview** |  |
| --- | --- | --- | --- | --- | --- | --- | --- | --- |
|  | N = 33 | % | N = 33 | % | N = 14 | % | N = 18 | % |
| **Symptoms** |  |  |  |  |  |  |  |  |
| Chest pain | 1 | 3 | 7 | 21 | 1 | 7 | 4 | 22 |
| Constitutional (fevers, tiredness, decreased appetite) | 3 | 9 | 11 | 33 | 2 | 14 | 5 | 28 |
| Gastrointestinal (nausea, vomiting, diarrhea, bloating, or abdominal pain) | 3 | 9 | 10 | 30 | 4 | 29 | 9 | 50 |
| Headache | 7 | 21 | 10 | 30 | 3 | 21 | 5 | 28 |
| Mental health (little interest, depressed, bipolar symptoms, anxiety, anger, panic, insomnia, or survivor's guilt) | 4 | 12 | 6 | 18 | 5 | 36 | 6 | 33 |
| Respiratory (SOB, pleuritic chest pain, or cough) | 4 | 12 | 17 | 52 | 7 | 50 | 4 | 22 |
| Other symptoms (hair loss, muscle atrophy, loss of singing voice, or other) | 0 | -- | 3 | 9 | 1 | 7 | 2 | 11 |
| **Health conditions<sup>1</sup>** |  |  |  |  |  |  |  |  |
| Behavioral (ADD, ADHD, Anxiety, Depression, PTSD, or substance use) | 16 | 49 | 9 | 27 | 2 | 14 | 0 | -- |
| Anxiety | 12 | 36 | 9 | 27 | 1 | 7 | 0 | -- |
| Depression | 8 | 24 | 5 | 15 | 0 | -- | 0 | -- |
| Other | 5 | 15 | 1 | 3 | 1 | 7 | 0 | -- |
| Cardiovascular (high blood pressure, cholesterol, or heart disease) | 4 | 12 | 3 | 9 | 2 | 14 | 1 | 6 |
| COVID-19 |  |  | 1 | 3 | 0 | -- | 1 | 6 |
| Diabetes | 1 | 3 | 1 | 3 | 0 | -- | 0 | -- |
| Respiratory (Sleep apnea, asthma, or COPD) | 9 | 27 | 4 | 12 | 1 | 7 | 0 | -- |
| Asthma | 9 | 27 | 3 | 9 | 1 | 7 | 0 | -- |
| Other (other conditions, arthritis, or chronic headaches) | 7 | 21 | 3 | 9 | 3 | 21 | 1 | 6 |
| **Health care utilization ≥1 visit** |  |  |  |  |  |  |  |  |
| Any healthcare provider |  |  | 30 | 91 | 13 | 93 | 15 | 83 |
| Cardiologist |  |  | 5 | 46 | 1 | 7 | 2 | 11 |
| Dentist |  |  | 2 | 6 | 1 | 7 | 1 | 6 |
| Mental health provider |  |  | 4 | 12 | 6 | 43 | 3 | 17 |
| Primary care provider |  |  | 18 | 55 | 4 | 29 | 9 | 50 |
| Pulmonologist |  |  | 27 | 79 | 2 | 14 | 1 | 6 |
| Urgent Care provider |  |  | 3 | 9 | 2 | 14 | 2 | 11 |
| Other health care provider |  |  | 3 | 9 | 6 | 43 | 6 | 33 |
| **Medication use** |  |  |  |  |  |  |  |  |
| Corticosteroid use |  |  | 23 | 70 | 0 | -- | 0 | -- |
| **Effects** |  |  |  |  |  |  |  |  |
| Help with personal care needs |  |  | 4 | 12 | 2 | 14 | 0 | -- |
| Help with routine needs |  |  | 9 | 27 | 1 | 7 | 2 | 11 |
| Financial problems |  |  | 10 | 30 | 0 | -- | 1 | 6 |

<sup>1</sup>Refers to that health condition being newly diagnosed in the period noted below for first interview, second interview, and third interview.
\*Refers to the time between EVALI hospitalization discharge (for those hospitalized) or diagnosis (for those not hospitalized) and first interview.
\*\*Refers to the 3month period before the interview.
Missing values were excluded from the table

### Symptoms

Between EVALI diagnosis and first interview, and during the three months before the second wave of interviews, respiratory symptoms were commonly reported (50%); however, gastrointestinal symptoms were most commonly reported (50%) during the three months before the third wave of interviews.

Among the 10 respondents who completed all three interviews, gastrointestinal symptoms were the most common (60%), and half (30%) remained unresolved by the third interview (Fig 2). Similarly, approximately half of respiratory, constitutional, and chest pain symptoms remained unresolved by the third interview.

**Fig 2.**
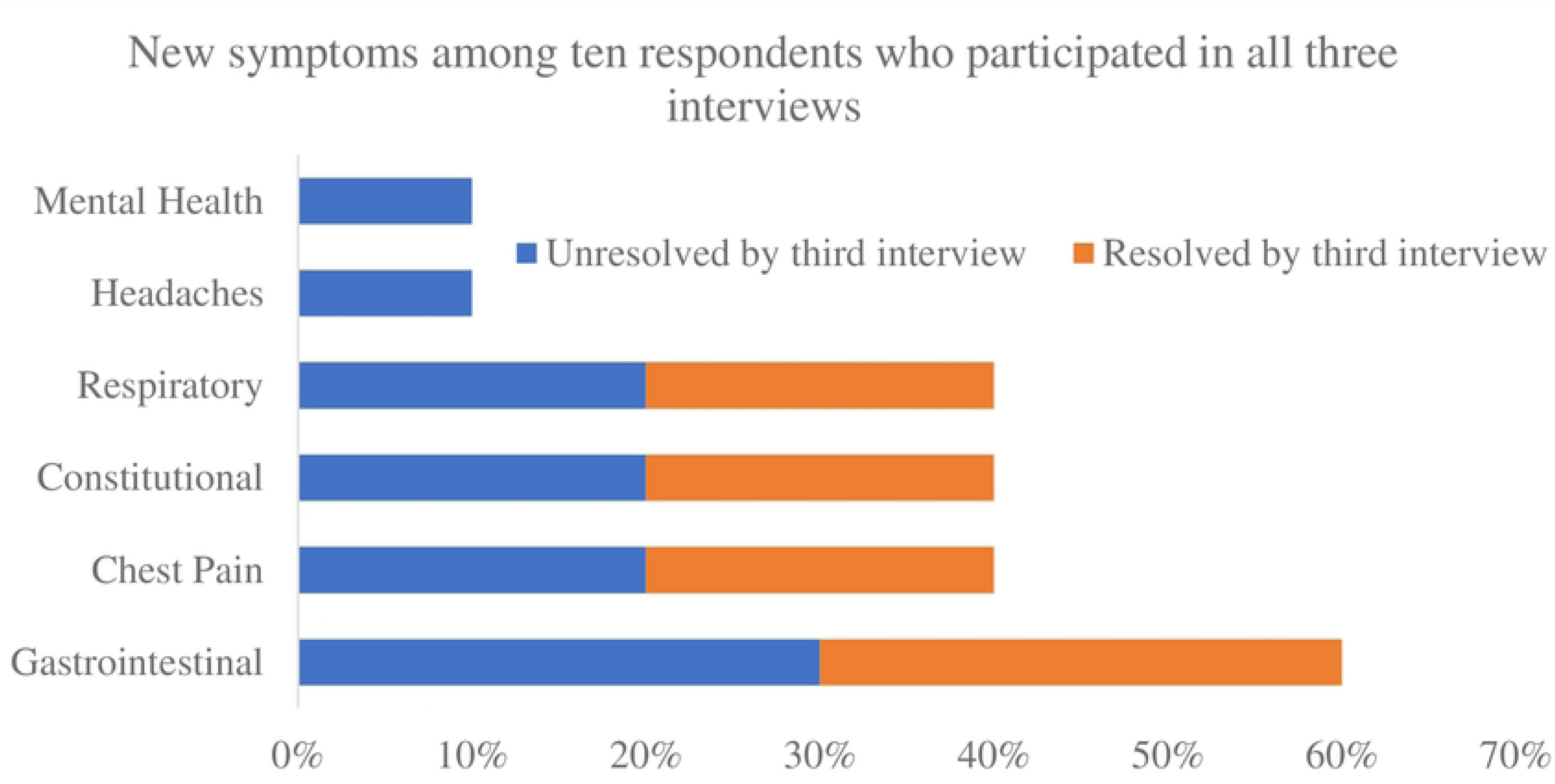
New symptoms after receiving EVALI diagnosis among the respondents who participated in all three interviews. *Mental health includes attention deficit disorder (ADD), attention deficit hyperactivity disorder (ADHD), anxiety, depression, or post-traumatic stress disorder (PTSD); respiratory includes sleep apnea, asthma, or chronic obstructive pulmonary disorder; constitutional includes fevers, tiredness, or decreased appetite; and gastrointestinal includes nausea, vomiting, diarrhea, bloating, or abdominal pain.

### Effects on personal function and finances

In the months after an EVALI diagnosis, nine of 33 respondents (27%) required help with routine needs such as everyday household chores, four (12%) required help with personal care such as bathing or eating, and 10 (30%) reported financial problems because of EVALI. By the third wave of interviews, only two (11%) respondents reported requiring help with routine needs, no respondents required help with personal care, and one (6%) respondent reported recent financial problems because of EVALI.

Most (70%, n = 23) respondents were employed and 33% (n = 11) were in school when diagnosed. All students returned to school or graduated, and 74% of those employed (n = 17) returned to the same job after their diagnosis. Nine (53%) of those who returned to the same job missed from one week to one month of work; 35% (n = 6) missed one week of work or less; and 6% (n = 1) missed from one to three months of work. Accommodations were made for 18% (n = 3) on return to work and 30% (n = 3) on return to school.

### Healthcare use

Between diagnosis or hospital discharge and first interview, 91% of respondents had received treatment by a health care provider, most often a pulmonologist (79%) or an ambulatory primary care provider (55%). Respondents continued to interact with health care providers, with 93% and 83% receiving treatment from healthcare providers during the three months preceding second and third interviews respectively; they most frequently visited mental health providers (43% and 17%) and primary care providers (29% and 50%) during this time frame.

### Respondent commentary on EVALI

Qualitative responses were received from 23 (70%) of the 33 study respondents, nine (40%) of whom were female. During the three waves of interviews, 14 (52%) respondents commented once, nine (39%) commented twice and two (8%) commented during all three interviews. This provided a total of 36 statements addressing current concerns, recovery, and long-term health outcomes (data not shown).

During the three interview waves, 10 of 23 respondents (43.5%) noted change in outlook about e-cigarette or vaping product use, specific product types, or health risks. Six respondents (26%) stated that they had stopped e-cigarette or vaping product use. However, three (13%) continued to report smoking cannabis, use of what they termed natural products or obtaining products from a medical cannabis dispensary. Eight (34.7%) respondents suggested they had recovered, five (22%) suggested they were still experiencing symptoms from injury or treatment. Four (18%) suggested the experience had been traumatizing and six (26%) expressed variable thoughts about the lack of effect their injury had on friends, lack of regulation, the need for provider education, or had no other significant concerns.

## Discussion

Respondents reported symptoms throughout the follow-up period, and approximately half reported new-onset morbidity. Although many symptoms resolved, especially during the latter part of the follow-up period, our qualitative findings indicate that changes to vaping behavior and mindset endured. The periodic interviews allowed us to collect data regarding symptoms, and effects on education and employment, although the COVID-19 pandemic likely played a role in some of the mental health symptoms and employment challenges reported.

CDC’s clinical guidance recommends outpatient primary care or pulmonary specialist follow up, optimally within 48 hours of discharge, and social support and access to mental health and substance use disorder services [12]. Although many of our respondents received a diagnosis before the October 2019 guidance release, overall treatment was consistent with this guidance.

Almost all respondents had been treated by ≥1 health care providers by their first interview. Per CDC, cardiac disease, chronic pulmonary disease, diabetes, and older age were risk factors for higher morbidity and mortality among those who received an EVALI diagnosis. Nationwide, anxiety, depression, attention deficit disorder (ADD), attention deficit hyperactivity disorder (ADHD), and other mental and behavioral health conditions were common among EVALI patients [2, 13]. Our findings were consistent with CDC’s nationwide findings; however, the mortality rate in NYC (7.5%) among those who received a diagnosis in 2019 was higher than the national rate (2.4%), which included cases through February 2020.

Approximately half (49%) of respondents had preexisting behavioral health conditions. The rate of preexisting depression in our cohort (24%) was approximately three times the overall rate in NYC (9%).[14] National survey data suggest that self-medication (e.g., use of alcohol, cannabis, or nicotine to address anxiety) for mental health conditions is not uncommon [15]. Approximately half of respondents reported new diagnoses for other physical or behavioral health conditions during the follow-up period. The possibility exists that some of these new diagnoses were preexisting, but undiagnosed conditions. Young adults generally use the health care system less than other age groups overall, but have higher rates of emergency department use [16]. Our findings indicate that closer medical follow-up after EVALI may have facilitated additional diagnoses and improved care. EVALI might have also been a contributing factor to some of the newly diagnosed behavioral health and respiratory conditions. Fewer than half of our respondents were treated by mental health providers. However, supporting recovery from EVALI will require some degree of mental health follow-up and substance use treatment to reduce the risk for reinjury or other secondary pulmonary complications. More consistent use of outpatient services for young adults might also prevent or reduce severity of future EVALI cases.

Two other published studies have looked at long-term outcomes after EVALI. Blagev et al tracked 73 patients in Utah through medical records abstraction and conducted clinical screening activities at 12-months post-diagnosis [17]. Triantafyllou et al retrospectively reviewed electronic medical records of 41 patients who received an EVALI diagnosis and admitted to any of the University of Pittsburgh Medical Center hospitals [18]. Both studies reported similar age and sex distributions, rates of behavioral health and cardiovascular disease, and proportions of respondents using e-cigarettes with THC or nicotine, consistent with our interview findings.

Blagev et al and our interviews reported similar median lengths of stay in the hospital (5 and 6 days respectively). Our findings contribute prospective data from a third jurisdiction at multiple timepoints on the physical, mental, and functional health of respondents, and social effects. One main difference between our findings and those from the other two studies is that Triantafyllou et al included hospital admissions through September 2020 and Blagev et al included diagnoses through August 2021. However, our respondents all received an EVALI diagnosis in 2019. Thus, our time frame excluded the potential for COVID-19 to be a factor in the initial hospitalization and case identification. In addition, compared with Triantafyllou et al, a higher proportion of our respondents reported a history of asthma (27% vs 8%). Among our respondents, although only some reported continued vaping (15%–29% across waves), most reported using non-vaped cannabinoids, often by smoking or dabbing (i.e., inhaling a smoked or aerosolized concentrated cannabis resin). Participants in the Blagev et al study were more likely to report nicotine consumption (vaped and smoked) and cannabis vaping after receiving their EVALI diagnosis than our respondents.

### Limitations

The limited number of potential respondents (n = 48) and the nonresponse rate (32%) suggests that the findings might not have captured the full range of post-discharge health sequelae among the potential respondents. However, respondents and nonrespondents did not differ by sex, age, initial hospitalization status, or length of stay. Economic or other circumstantial differences imposed by the COVID-19 pandemic might have been a barrier to participation or with ongoing follow-up. The conclusions also involved self-reported findings from a local municipal population, which cannot be generalized to the broader population affected by EVALI, especially given the variable state and local regulatory environment for cannabis and e-cigarettes. A robust effort was made to abstract race and ethnicity information from available electronic health records; however, these data were found to be incomplete and inadequate for identifying associated disparities [19]. Although the interviews were confidential and used nonjudgmental survey questions administered by experienced interviewers, they were not anonymous. The possibility exists that some persons responded to the surveys based on perceptions of social desirability, need to share their story for benefit of others, or to affirm a positive outcome with their health. Because of DOHMH staffing constraints during the pandemic, some respondents also did not speak with the same investigator each time.

### Public Health Implications

Limited regulation and oversight of new consumer products, including ingredients in nicotine and cannabis products, contributed to the EVALI outbreak.[20] These products continue to evolve, which presents a challenge to the development and communication of health risk information. Ongoing surveillance is particularly crucial, because cannabis was legalized for adult use in NYS in 2021, and new products continue to emerge. Although vitamin E acetate has been identified as a contributor to EVALI, certain respondents vaped only nicotine products before receiving a diagnosis, which are not known to contain vitamin E acetate. Consistent with the conclusion reached by other researchers, we lack sufficient information about vaping product constituents and excipients involved in each lung injury to provide specific new warning to the public or establish more protective regulations for manufacturers.^1^

Given the rapid evolution of products, periodic follow-up with a group of consumers might help supplement other traditional modes of surveillance. For example, one respondent inquired about risks for nicotine-free vaping products being marketed as wellness products or personal diffusers, which claimed to deliver essential oils. This led NYC DOHMH to update communications for the public, retailers, and clinicians to include these products.

NYC DOHMH maintains some active and passive surveillance of EVALI, identifying approximately three or four cases of EVALI per month. As the COVID-19 pandemic progressed during spring of 2020, we noted that the clinical assessment of patients began to evolve, often with repeated COVID-19 testing, which highlighted the continued challenge of recognizing and diagnosing EVALI. Detailed substance use histories often led to the ultimate diagnosis and appropriate treatment. Subsequently, state and local agencies collaborated to develop guidance informing health care providers that EVALI cases continue to occur and should be considered alongside other etiologies.[21]

The preponderance of preexisting and new mental health diagnoses among respondents also suggests that clinical follow up by behavioral health providers might be beneficial for addressing substance use and psychological trauma of injury, hospitalization, and any emotional and social adjustments to long-term physical impairments suffered by patients.

### Conclusion

NYC residents with EVALI reported symptoms throughout the follow-up period, and approximately half reported new-onset morbidity. These findings have guided clinician education and public health messaging and might influence e-cigarette policy and individual health behaviors. Findings and follow-up have also guided surveillance efforts, both for other emerging products and for emerging injuries or illnesses that might result in chronic disease. Other studies are needed to understand long-term health of EVALI cases and to more fully understand causes of EVALI associated with various vaping products.

## Data Availability

Raw data used for producing this report are not available because they were collected by a Public Health Authority as public health surveillance and not research as defined by (45 CFR 46.102(l)(2)). The small cohort makes protecting confidentiality particularly important from an ethical standpoint. However, we believe publishing these findings, without making the data available, will promote more effective protection and promotion of public health.

## Acknowledgments

We wish to acknowledge Tabassum Z. Insaf, PhD, Monica P. Nordstrom, MSPH, from the New York State Department of Health, and Philip C. DiSalvo, MD, of the Carle Illinois College of Medicine, formerly of the New York City Department of Health’s Poison Control Center and the staff of the Poison Control Center, for their exceptional assistance supporting our lung injury surveillance and identification of New York City EVALI cases in 2019. We also acknowledge Kim Kessler, JD, Assistant Commissioner of the Bureau of Chronic Disease Prevention (BCDP), New York City Department of Health and Jennifer G. Wright, DVM, MPH from the Centers for Disease Control and Prevention’s Division of Partnership Support (PHIC) and Division of Workforce Development (DWD) for their detailed manuscript reviews, edits and comments. We also thank Lourdes M. Cifuentes, MPH, of the Food Bank for New York City, a former BCDP staff member, who conducted interviews and supported the data collection process during the first wave of interviews.

## Footnotes

1 45 C.F.R. part 46, 21 C.F.R. part 56; 42 U.S.C. Sect. 241(d); 5 U.S.C. Sect. 552a; 44 U.S.C. Sect. 3501 et seq

